# High admission blood glucose is an independent risk factor of poor prognosis in COVID-19: A systematic review and dose-response meta-analysis

**DOI:** 10.1101/2020.09.25.20200774

**Authors:** Gilbert Lazarus, Jessica Audrey, Vincent Kharisma Wangsaputra, Alice Tamara, Dicky L. Tahapary

## Abstract

**Aims:** To investigate the prognostic value of admission blood glucose (BG) in predicting COVID-19 outcomes, including poor composite outcomes (mortality/severity), mortality, and severity.

**Materials and methods:** Eligible studies evaluating the prognostic value of fasting BG (FBG) and random BG (RBG) levels in predicting COVID-19 outcomes were included and assessed for risk of bias with the Quality in Prognosis Studies tool. Random-effects high-vs-low meta-analysis followed by dose-response analysis using generalized least squares model in a two-stage random-effects meta-analysis were conducted. Potential non-linear association was explored using restricted cubic splines and pooled using restricted maximum likelihood model in a multivariate meta-analysis.

**Results:** The search yielded 35 studies involving a total of 14,502 patients. We discovered independent association between admission FBG and poor prognosis in COVID-19 patients. Furthermore, we demonstrated non-linear relationship between admission FBG and severity (P_non-linearity_<0.001), where each 1 mmol/L increase augmented the risk of COVID-19 severity by 33% (risk ratio 1.33 [95% CI: 1.26-1.40]). Albeit exhibiting similar trends, study scarcity limited the strength of evidence on the independent prognostic value of admission RBG. GRADE assessment yielded high-quality evidence for the association between admission FBG and COVID-19 severity, and moderate-quality evidence for its association with mortality and poor outcomes, while the other assessments yielded very low-to-low quality.

**Conclusion:** High level of FBG at admission was independently associated with poor prognosis in COVID-19 patients. Further researches to confirm the observed prognostic value of admission RBG and to ascertain the estimated dose-response risk between admission FBG and on COVID-19 severity are required.

## INTRODUCTION

The rapidly spreading coronavirus disease 2019 (COVID-19) has placed significant burdens on healthcare systems worldwide, with millions of cases and hundred-thousands of deaths.^1^ Despite significant efforts in comprehending the disease, its diagnosis and prognostication remain challenging, attributing to the prevalent non-specific and atypical symptoms.^2^ In light of this, recent reports have indicated that admission blood glucose (BG) may yield prognostic values in predicting COVID-19 outcomes.^3,4^ Nevertheless, to the best of our knowledge, no meta-analysis has evaluated the prognostic value of admission BG level in predicting the outcomes of COVID-19 patients. Hence, this review intends to summarize the current knowledge regarding the role of admission BG level in determining the prognosis of COVID-19 patients.

## MATERIALS AND METHODS

This systematic review was conducted in accordance with the guideline recommended by the Cochrane Prognosis Methods Group^5^ and reported based on the Preferred Reporting Items for Systematic Reviews and Meta-Analyses (PRISMA) statement^6^. A detailed protocol has been previously registered in PROSPERO (CRD42020154772^7^).

### Search strategy

Two independent investigators performed thorough literature searches, with discrepancies resolved by a third investigator in a blinded fashion. Searches were conducted through PubMed, Ovid EMBASE, CENTRAL, EBSCO MEDLINE, CINAHL, Scopus, and WHO COVID-19 database for studies published up to 8 September 2020. Grey literature (e.g., preprints) search and literature snowballing of references were also performed. No language restrictions were applied. Details on databases and keywords are further elaborated in **Appendix Table S1**.

### Study eligibility criteria

Inclusion criteria were set to filter primary studies investigating the association between admission BG level and poor outcomes among COVID-19 patients (see **Appendix Table S2**). Admission BG level was determined from the first BG measurement following patients’admission to hospital prior to any intervention, while poor outcomes were further dichotomized into mortality and severity.^8^ Conversely, studies were excluded if any of the following criteria were met: (1) case reports, case series, or letter to editors; (2) irretrievable full-text articles; or (3) non-English articles.

### Data extraction and risk of bias assessment

Data extraction and risk of bias assessment were performed by two independent reviewers using a pre-specified form, with discrepancies resolved by the consensus with an independent third investigator. Details on the data items and handling are further discussed in **Appendix pg**.**7**. The main outcome of interest in this review was the risk of poor composite outcomes (i.e. mortality/severity), mortality, and severity among COVID-19 patients. Whenever possible, outcomes on severity were further investigated per following criterion, including invasive ventilation, intensive care unit (ICU) admission, acute respiratory distress syndrome (ARDS), and shock.^8^ Included studies were further assessed for methodological quality by using the Quality in Prognosis Studies tool^9^ and subsequently judged to be yielding low, moderate, or high risk of bias **(Appendix Table S3)**. Lastly, the certainty of the evidence was evaluated using the modified Grading of Recommendations Assessment, Development, and Evaluation (GRADE) framework, where the quality of evidence was regarded as high, moderate, low, or very low.^10^

### Statistical analysis

Analyses were performed for both adjusted and unadjusted estimates; however, adjusted estimates were primarily utilized for reporting and interpretation of results.^11^ Pooled effects were converted to and presented in risk ratios (RRs; see **Appendix pg. 11**). Quantitative synthesis was first conducted by comparing the highest vs lowest categories of exposures by using the generic inverse variance method with the DerSimonian-Laird random-effects model^12^. Whenever appropriate (n≥10), potential publication bias was evaluated visually by contour-enhanced funnel plot^13^ and quantitatively by Egger’s^14^ and Begg’s^15^ tests. Statistical heterogeneity was investigated with Cochran’s Q test and I^2^ statistics. Dose-response meta-analysis (DRMA) was conducted only for adjusted outcomes. Study-specific linear trend was estimated using the generalized least squares method and pooled using the two-stage random effects meta-analysis. Meanwhile, potential non-linear dose-response trend was evaluated using restricted cubic splines with three-knots model at the 10^th^, 50^th^, and 90^th^ percentiles, and subsequently pooled using the restricted maximum likelihood method in a multivariate random-effects meta-analysis. The Wald test was used to assess for non-linearity.

A priori, we determined subgroup and sensitivity analyses only for adjusted results. Whenever available, subgroup analyses were carried out based on study design, location, sample size, risk of bias, number of categories, effect size type, and diabetic status. On the other hand, sensitivity analyses were conducted by leave-one-out analysis and the exclusion of studies with high-risk of bias. For DRMA, subset analysis was performed according to diabetic subgroups, while sensitivity analyses were conducted by assigning alternative approaches for open-ended categories (for linear trends) and alternative knots locations (for non-linear trends). Meta-analysis was conducted with R ver. 4.0.0 (R Foundation for Statistical Computing, Vienna, Austria)^16^, and additional analyses with MetaXL software ver.5.3. (EpiGear International, Queensland, Australia)^17^. The significance level was set at 5% for all analyses. Further details on DRMA and additional analyses are discussed in **Appendix pg. 12-13**.

## RESULTS

### Study selection and characteristics

The details on the literature search process are summarized on **Figure 1**. The initial search yielded 1177 articles, of which 636 were deduplicated and 482 were excluded following title and abstracts screening, resulting in the retrieval of 59 records for full-text assessments–among which 13 inappropriate design, five inappropriate settings, four incompatible language, two irretrievable full-text articles, and one unidentifiable setting (**see Appendix pg. 12-13** for further details) were excluded. Consequently, a total of 35 studies with 14,502 patients were included in this systematic review, where 7918 patients were male (54.6%), and hypertension (4940 [34.1%]) as well as diabetes (4540 [31.3%]) were the most reported comorbidities **(Appendix Table S4**). In quantitative analysis, 10 studies were excluded as seven^18–24^ only reported P-value and three^25–27^ reported different effect measures.

**Figure 1.**
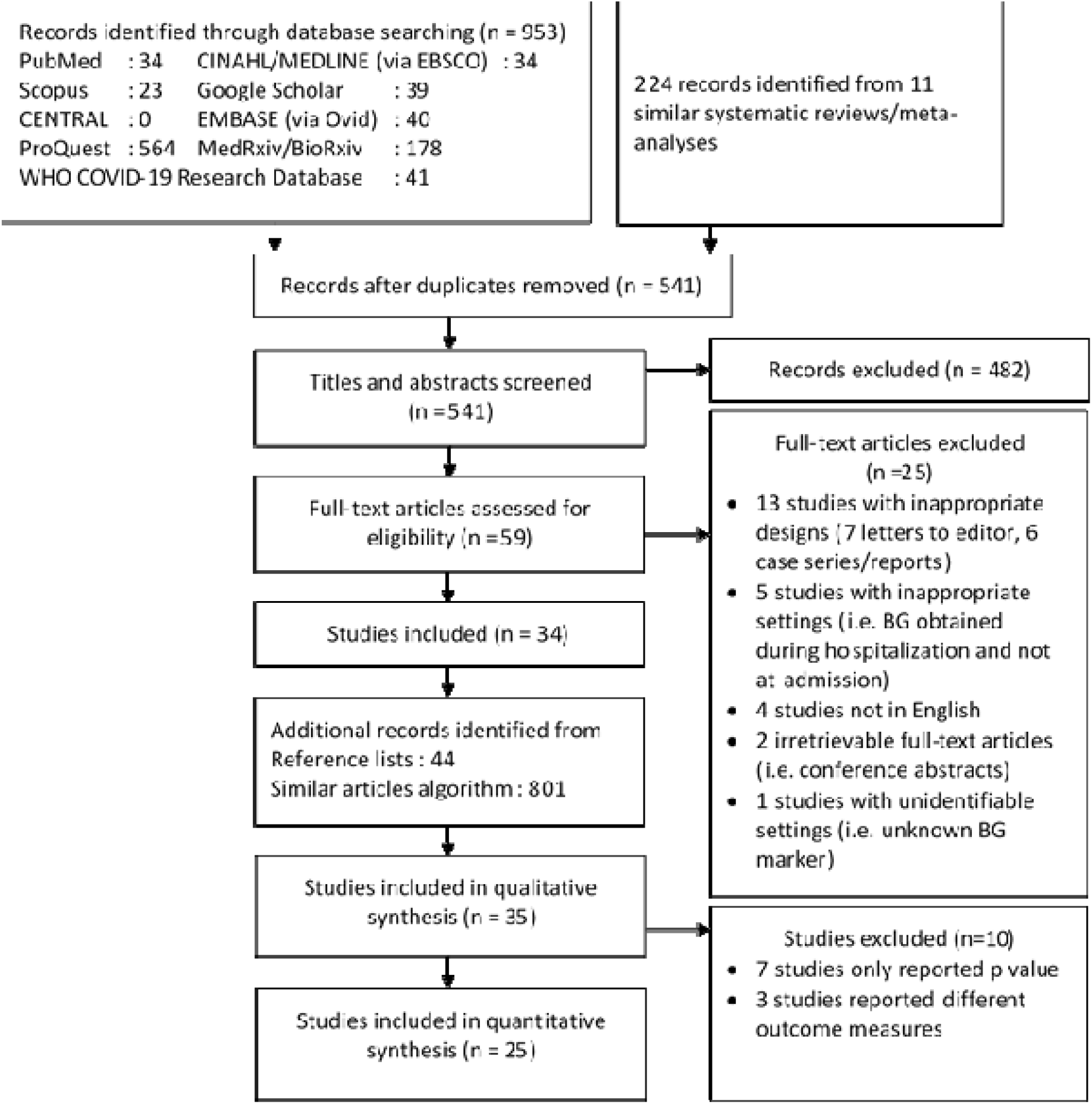
Diagram flow illustrating the literature search process and results. CENTRAL, Cochrane Central Register of Controlled Trials; CINAHL, Cumulative Index to Nursing and Allied Health Literature. WHO, World Health Organization.

From 35 included studies, more than half were conducted in China (23 studies), while the others were five each in America^18,21,27–29^ and Europe^20,25,30–32^, and one each in Hong Kong^33^ and South Korea^34^. FBG was utilized in 23 studies, while RBG in 13 studies. Bias assessment revealed a predominant low-to-moderate risk of bias (16 and 12 studies, respectively). Most of the studies yielded unclear risk of bias in study participation and confounding domains (**Figure 2 and Appendix Figure S1**), which may partly be explained by the fact that all but two studies^21,29^ were conducted retrospectively.

**Figure 2.**
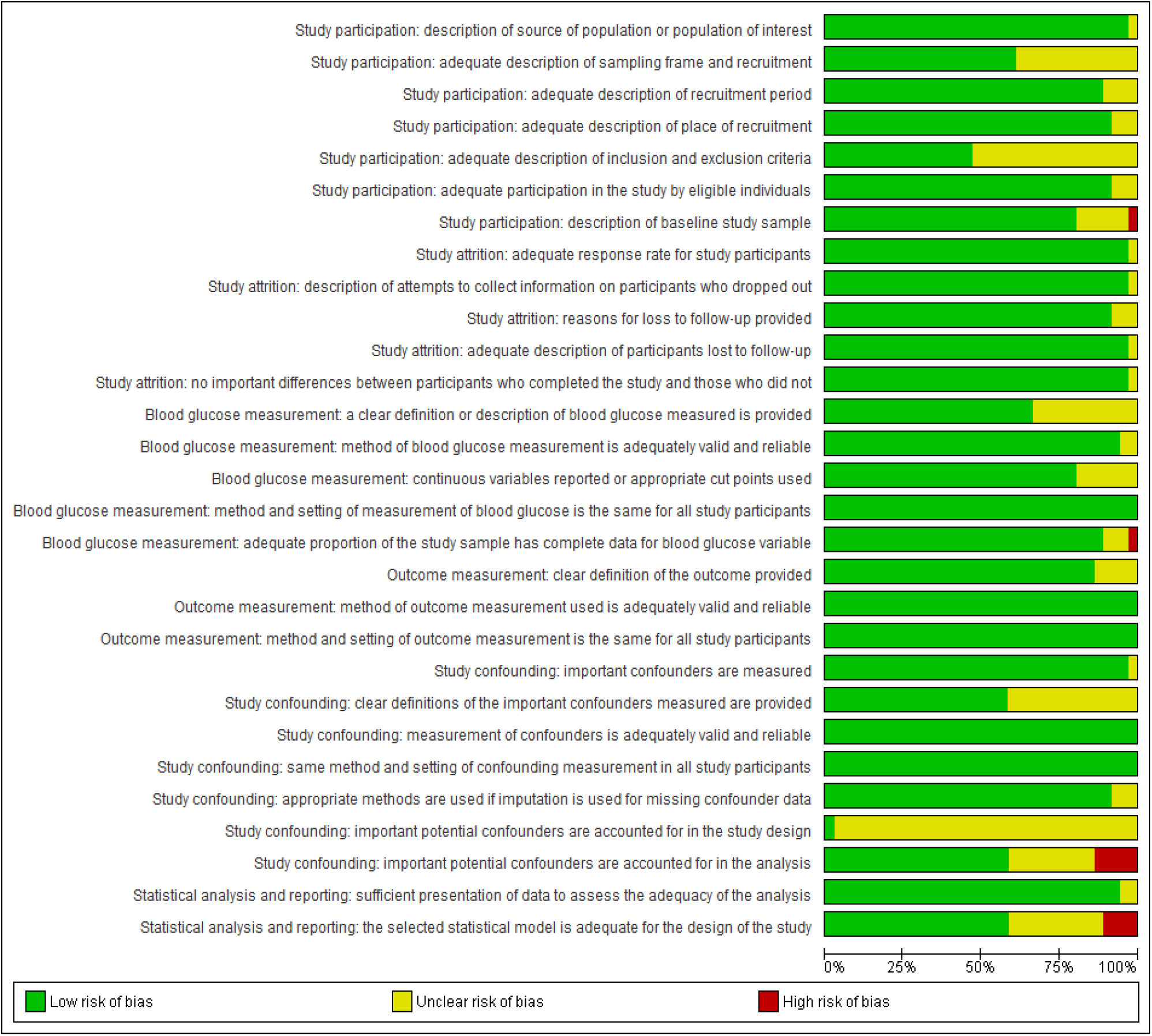
Risk of bias graph summarizing the results of each risk of bias item in percentages across all studies.

### Outcomes

The summary of adjusted and unadjusted pooled effects of high vs low meta-analysis are consecutively listed in **Table 1** and **Appendix Table S5**; and the certainty of evidence as assessed with the GRADE approach is summarized in **Appendix Table S6**. GRADE assessments of the prognostic value of FBG resulted in high-quality evidence for severity and moderate-quality evidence for mortality and poor outcome, whereas the remaining domains yielded very low-to-low-quality evidence. Overlapping populations were observed in four studies^35–38^ **(Appendix Table S7)**, and analyses were prioritized to Wang et al.^35^ due to larger sample size.

**Table 1.** Summary of high versus low meta-analysis and subgroup analyses

| Outcomes | Studies | Events/N | RR (95% CI) | Heterogeneity |  |
| --- | --- | --- | --- | --- | --- |
|  |  |  |  | I<sup>2</sup> | P value |
| **Fasting blood glucose<sup>†,‡</sup>** |  |  |  |  |  |
| Poor composite outcome | 4 | 429/1184 | 1.20 (1.04-1.39) | 84% | <0.001 |
| Diabetic status |  |  |  |  |  |
| Diabetic | 2 | 108/184 | 1.10 (1.03-1.17) | 0% | 0.562 |
| Non-diabetic | 3 | 367/848 | 1.75 (1.02-2.99) | 95% | <0.001 |
| Mortality<sup>§</sup> | 10 | 827/3814 | 1.81 (1.41-2.33) | 87% | <0.001 |
| *Subgroup analysis<sup>‡</sup>* |  |  |  |  |  |
| Categories |  |  |  |  |  |
| Two categories | 6 | 602/2494 | 2.26 (1.75-2.91) | 2% | 0.404 |
| 11.1 mmol/L | 1 | 19/132 | 5.66 (1.36-12.89) | NA | NA |
| 7.0 mmol/L | 3 | 167/1284 | 2.60 (1.72-3.93) | 0% | 0.608 |
| 6.1 mmol/L | 2 | 416/1078 | 1.68 (1.25-2.26) | 0% | 0.640 |
| More than two categories | 2 | 147/917 | 3.24 (1.28-8.19) | 58% | 0.121 |
| Continuous | 2 | 78/403 | 1.02 (1.01-1.02) | 0% | 0.842 |
| Sample size |  |  |  |  |  |
| <200 patients | 3 | 65/307 | 2.52 (0.73-8.66) | 86% | 0.001 |
| ≥200 patients | 7 | 762/3507 | 2.05 (1.34-3.15) | 85% | <0.001 |
| Risk of bias |  |  |  |  |  |
| Low | 8 | 700/2767 | 2.38 (1.50-3.78) | 85% | <0.001 |
| Moderate/High | 2 | 127/1047 | 1.37 (0.68-2.77) | 82% | 0.019 |
| ES type |  |  |  |  |  |
| RR | 4 | 386/1225 | 2.10 (1.08-4.10) | 87% | <0.001 |
| HR | 6 | 441/2589 | 2.13 (1.25-3.61) | 87% | <0.001 |
| Diabetes status |  |  |  |  |  |
| Diabetic | 4 | 245/586 | 1.70 (0.91-3.19) | 70% | 0.018 |
| Non-diabetic | 6 | 605/2018 | 2.10 (1.16-3.79) | 90% | <0.001 |
| Severity<sup>§</sup> | 7 | 824/3280 | 3.65 (2.31-5.75) | 78% | <0.001 |
| *Subgroup analysis<sup>‡</sup>* |  |  |  |  |  |
| Categories |  |  |  |  |  |
| Two categories | 3 | 423/1891 | 2.31 (1.55-3.46) | 21% | 0.281 |
| More than two categories | 4 | 401/1389 | 5.64 (2.35-13.54) | 88% | <0.001 |
| Sample size |  |  |  |  |  |
| <200 patients | 2 | 77/201 | 2.85 (1.63-4.98) | 13% | 0.284 |
| ≥200 patients | 5 | 747/3079 | 4.08 (2.21-7.52) | 85% | <0.001 |
| Risk of bias |  |  |  |  |  |
| Low | 5 | 707/2808 | 2.30 (1.98-2.68) | 3% | 0.391 |
| Moderate/High | 2 | 117/472 | 15.04 (7.03-32.19) | 0% | 0.368 |
| ES type |  |  |  |  |  |
| RR | 5 | 431/1278 | 4.99 (2.41-10.34) | 84% | <0.001 |
| HR | 2 | 393/2002 | 2.40 (1.37-4.20) | 56% | 0.131 |
| Diabetes status |  |  |  |  |  |
| Diabetic | 2 | 112/230 | 1.58 (0.74-3.37) | 87% | 0.005 |
| Non-diabetic | 5 | 963/2936 | 2.67 (1.52-4.71) | 97% | <0.001 |
| **Random blood glucose** |  |  |  |  |  |
| ICU admission | 2 | 120/423 | 1.25 (0.84-1.84) | 86% | 0.008 |

<sup>†</sup>Subgroup analyses to explore potential source of heterogeneity was not possible as no subgroup yielded a minimum of two studies. <sup>‡</sup>Subgroup analysis based on study design and location were not possible as all included studies were retrospective and conducted in China. <sup>§</sup>Overlapping populations were observed between Li et al.<sup>38</sup>, Wang et al.<sup>35</sup>, Zhang et al.<sup>36</sup>, and Zhu et al.<sup>37</sup>, of which Wang et al.<sup>35</sup> was prioritized for analysis due to larger sample size.
**COVID-19**, Coronavirus disease 2019; **ICU**, intensive care unit

### Fasting blood glucose

We demonstrated that FBG was independently associated with poor prognosis in COVID-19 patients (**Table 1** and **Figure 3A-C**), although all models yielded substantial heterogeneity (I^2^=84% for poor outcome; I^2^=87% for mortality, I^2^=78% for severity; all with P_heterogeneity_<0.001). Subgroup analyses based on study design and location were not possible as all studies were retrospective and conducted in China. Furthermore, publication bias assessment was only eligible for mortality outcome.

**Figure 3.**
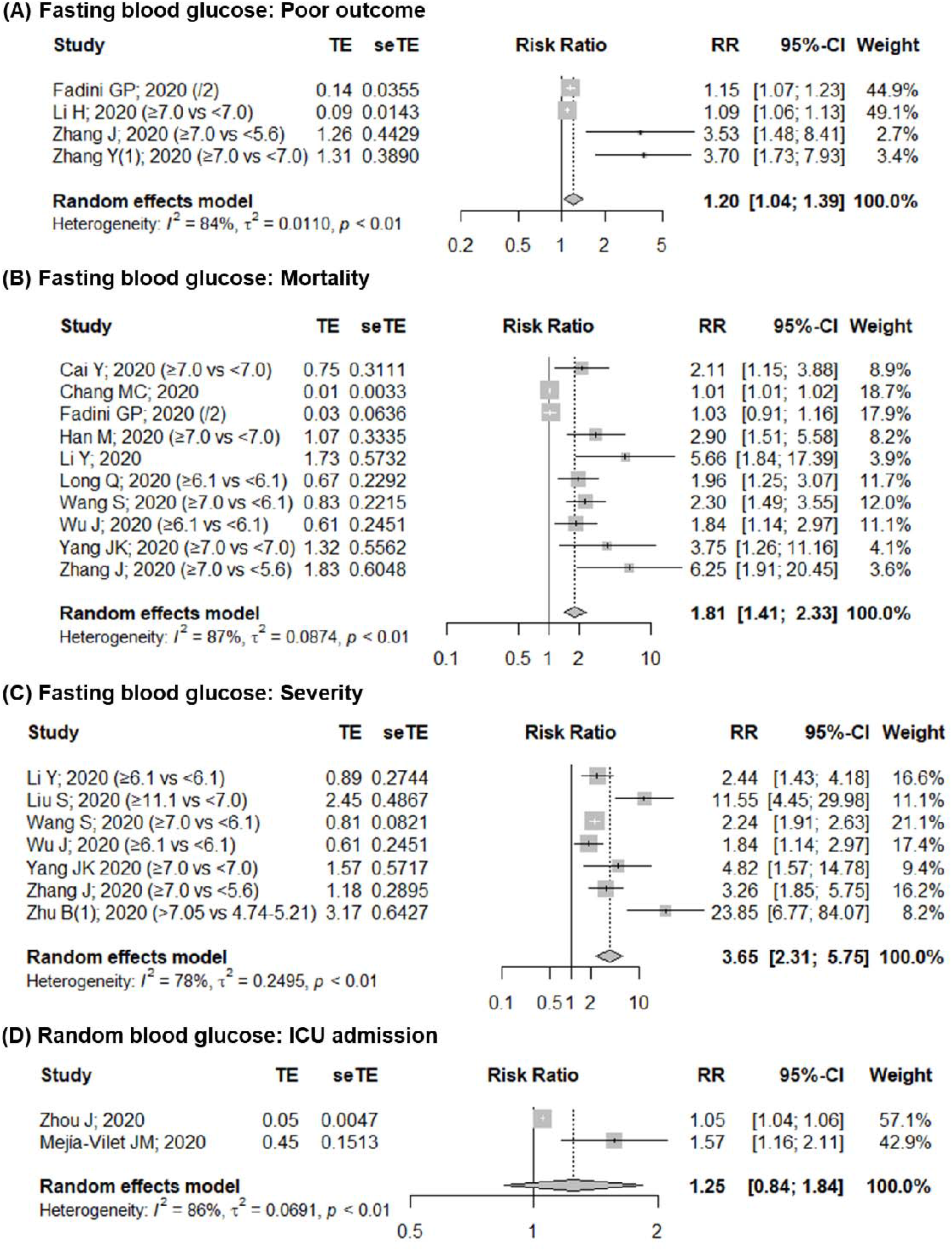
Pooled adjusted effects of high vs low meta-analysis comparing the association between (A) admission FBG and poor composite outcome, (B) admission FBG and mortality, (C) admission FBG and severity, and (D) admission RBG and ICU admission. **FBG**, fasting blood glucose; **ICU**, intensive care unit; **RBG**, random blood glucose.

We discovered that high admission FBG increased the risk of poor outcomes by 20% (RR 1.20 [95% CI: 1.04-1.39]; **Figure 3A**). However, we were unable to establish firm evidence as the observed heterogeneity remained unexplained (see footnote in **Table 1**) and the observed effects were diminished following the exclusion of Fadini et al.^32^ or Li et al.^38^ **(Appendix Figure S2A)**. In DRMA comprising of two studies^32,39^, we failed to observe exposure-response gradient (RR 1.23 [95% CI: 0.90-1.68]; P_heterogeneity_=0.008; **Figure 4A**), although study-specific slopes indicated that such trend exists. Considering this, we deemed the quality of evidence to be moderate for qualitative assessment and low for quantitative assessment.

**Figure 4.**
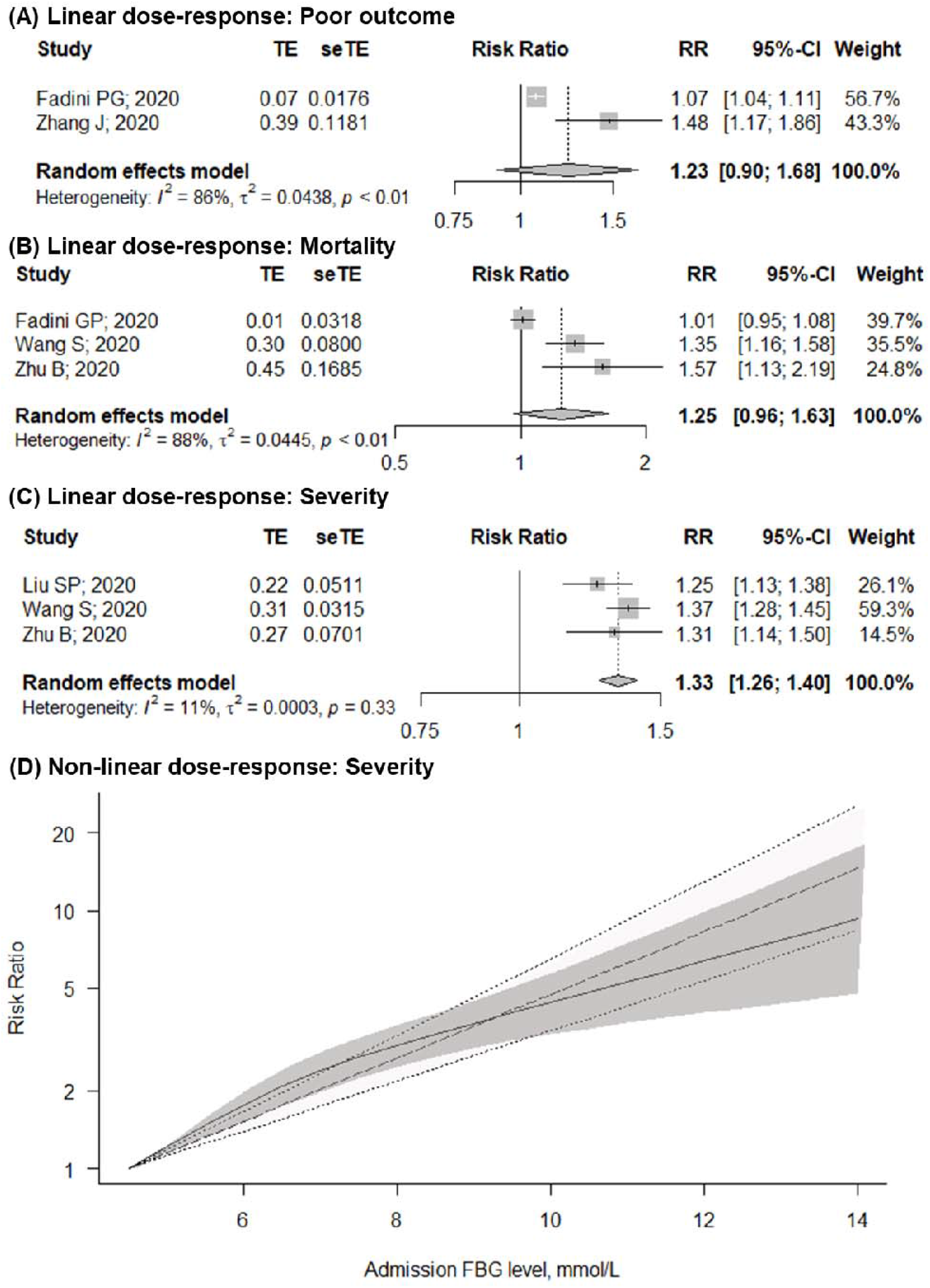
Results of dose-response meta-analysis illustrating linear trend between admission FBG and (A) poor outcome, (B) mortality, and (C) severity; and (D) non-linear trend between admission FBG and severity. In Figure C, darker area represents 95% confidence interval of non-linear trend, while lighter area represents 95% confidence interval of linear trend. **FBG**, fasting blood glucose.

For outcomes on mortality, we found a relatively consistent result (RR 1.81 [95% CI: 1.41-2.33]; **Figure 3B**); although the observed heterogeneity remained unexplained following subgroup analysis, except when the analysis was stratified according to its category (i.e. I^2^=2% for studies reporting two categories). Furthermore, we discovered that the smaller-sized studies yielded excessively wide CIs as compared to larger studies **(Table 1)**. Nonetheless, sensitivity analysis suggested that these small studies did not contribute much to the pooled estimate as our findings remained consistent, except when Fadini et al.^32^ or Chang et al.^34^ were excluded, which exaggerated the pooled estimates **(Appendix Figure S2B)**. We detected asymmetry in the funnel plot which was further ascertained by Egger’s test (P<0.001; **Appendix Figure S3**), although Begg’s test indicated otherwise (P=0.245), suggesting the presence of publication bias. Nevertheless, we did not downgrade the outcome for publication bias upon qualitative assessment as most studies adequately adjusted for potential confounders **(Appendix Table S8)**.

DRMA for mortality outcome included three studies^32,35,39^ as overlapping populations were detected in two studies.^35,38^ We were also unable to observe any exposure-response gradients although most studies reported otherwise^35,38,39^. However, when analysis was conducted only for non-diabetic patients, we observed an elevated mortality risk of about 38% per 1 mmol/L increase (RR 1.38 [95% CI: 1.21-1.57]; **Appendix Figure S4**). Despite this, we decided to upgrade the certainty of evidence for exposure-response gradient only for qualitative summary, resulting in the judgment of evidence quality for qualitative and quantitative analysis to be moderate and very low, respectively. This implied that we were confident that admission FBG was independently associated with mortality in COVID-19 patients, but the interpretation of the pooled estimate should be made with caution. Non-linear DRMA for poor outcome and mortality were not conducted since only one study reported three or more categories for each outcome.^35,39^

With regards to outcomes on severity, we revealed that high admission FBG level increased the patient’s risk of developing severe COVID-19 infection by more than three-folds (RR 3.65 [95% CI: 2.31-5.75]). Although considerable heterogeneity was observed (I^2^=78%, P_heterogeneity_<0.001), subgroup analysis according to risk of bias explained all heterogeneity **(Table 1)**, suggesting that studies with moderate-to-high risk of bias exaggerated the estimates. Nevertheless, sensitivity analysis revealed similar estimates **(Appendix Figure S2C)**, suggesting that the pooled effects were mainly derived from studies with low bias risk.

DRMA for severity outcome consisted of three studies^35,40,41^ cumulating a total of 1077 patients. We discovered that each 1 mmol/L increase in admission FBG level increased the risks of developing severe COVID-19 infection by 33% (RR 1.33 [95% CI: 1.26-1.40]**; Figure 4B**). Moreover, we observed a non-linear relationship (P_non-linearity_<0.001), where the risks of progression to severe COVID-19 cases decelerated starting from 6.6 mmol/L and re-exponentiated starting from admission FBG level of 8.1 mmol/L **(Figure 4C)**. Both linear and non-linear trends remained consistent following sensitivity analyses **(Appendix Table S8 and Figure S5)**. When dose-response analysis was conducted only for non-diabetic patients, we observed a slightly higher risk (RR 1.36 [95% CI: 1.28-1.44]; **Appendix Table S9**).

The observed effects were more accentuated in patients without history of diabetes, where high admission FBG level increased the risk of poor outcome for diabetic patients by 10% and non-diabetic patients by a whopping 75% (diabetic vs non-diabetic; RR 1.10 [95% CI: 1.03-1.17] vs 1.75 [1.02-2.99]). Furthermore, diabetes-stratified analyses for mortality and severity outcomes revealed that the observed trends were non-significant for diabetic subgroup, whereas the models for non-diabetic subgroups remained robust **(Table 1)**.

Due to the paucity of studies reporting specific criteria of COVID-19 severity (i.e. ICU admission, invasive ventilation, ARDS, shock), we were unable to ascertain the independent effects to the respective outcomes. Nonetheless, the results were coherent to the main outcomes as **t**he pooled unadjusted effects showed similar trends in predicting ICU admission, invasive ventilation, ARDS, and shock **(Appendix Table S5)**. However, as our findings were limited, we judged the certainty of evidence to be low for ICU admission and invasive ventilation, and very low for ARDS and shock **(Appendix Table S6)**.

### Random blood glucose

Similar to FBG, we discovered that COVID-19 patients with high RBG level at admission were more susceptible to poor prognosis **(Appendix Table S5)**. Nevertheless, we were unable to establish a strong evidence on the independent prognostic value of admission RBG due to paucity of studies and equivocal trends. High vs low meta-analysis for adjusted effects was only eligible for outcomes on ICU admission, which resulted in non-significant estimate (RR 1.25[95% CI: 0.84-1.84]; **Figure 4D**), although study-specific estimates suggested otherwise. Considering this, we judged the quality of evidence to be low for mortality, and very low for the remaining outcomes **(Appendix Table S6)**.

With regards to mortality, we found that the prognostic value of admission RBG was independent of age and sex.^25,31^. However, Cariou et al. reported that the observed effect diminished following adjustments for clinical and biological features (odds ratio 1.30 [95% CI: 0.94-1.82]),^25^ while Coppelli et al. stated otherwise,^31^ Moreover, exposure-response trend were also observed in two studies,^31,42^ where Coppelli et al. reported that risk of mortality increased across quintiles of admission RBG (Q4 vs Q1, hazard ratios [HR] 5.91 [95% CI: 1.73-20.19]) and reached threshold effect at the highest quintile (Q5 vs Q1, HR 1.70 [95% CI: 0.49-5.90]; **Appendix Table S10**).^31^ However, we were unable to perform a formal dose-response analysis due to insufficient information. In addition, two studies also reported that the risk of mortality was more accentuated in patients without history of diabetes^30,31^, as ascertained by Coppelli et al.– reporting that the hazard of mortality was more robust in non-diabetic than in diabetic patients. (vs. normoglycemia; hyperglycemia: HR 2.39 [95% CI: 1.10-5.19], diabetes: HR 0.78 [95% CI: 0.29-2.09]).^31^ Subgroup and sensitivity analyses, as well as publication bias assessment and DRMA was not conducted due to insufficient number of studies.

## DISCUSSION

This meta-analysis showed that high admission BG level was associated with poor prognosis in COVID-19 patients. Although we were unable to establish a firm evidence on the independent prognostic value of admission RBG, our results on admission FBG level was consistent and robust. Furthermore, we also demonstrated dose-response trend between admission FBG level and COVID-19 severity. Although the potential non-linear association between admission FBG level with poor composite and mortality outcomes remained unexplored due to paucity of studies, we were able to establish non-linear relationship between admission FBG level and severity. However, in contrast with Zhu et al.^37^, we did not observe a J-shaped association between admission FBG and COVID-19 severity, which may be explained by the fact that all but one study^37^ reported only three categories.

Further analysis indicated that the observed effects were more accentuated in patients without prior history of diabetes; which was intriguing, considering the fact that there was as increasing proportion of COVID-19 patients presenting with hyperglycemia despite having no prior diabetic history.^32,35,43^ Furthermore, our findings indicated that at-admission hyperglycemia was associated with poorer outcomes regardless of prior diabetes status, suggesting the existence of a more direct link between glycemic status and poor COVID-19 outcomes.

The relationship between COVID-19 severity and hyperglycemia is possibly bidirectional, wherein infection might bring about state of stress and trigger an enhanced release of pro-inflammatory cytokines which may lead to insulin resistance.^44^ Stress may also induce the release of stress hormones which trigger liver glycogenolysis, aggravating the effects.^45^ Furthermore, SARS-CoV-2 is known to bind to angiotensin-converting enzyme 2 (ACE2) receptors, which are found to be expressed in pancreatic beta-cells, thus rendering it a target for the viral attack. Such binding provides a route for the virus to enter and damage the pancreatic islets, resulting in a defect of insulin production, as indicated in previous study with its SARS virus counterpart.^46^ Together, these factors may contribute to the development of acute hyperglycemia in COVID-19 patients.

The mechanism by which acute hyperglycemia drives the progression of COVID-19 remains largely unexplored. A study by Fadini et al. found that a decline in respiratory parameters was most responsible for mediating the effects of hyperglycemia on the outcome.^32^ Diabetes and hyperglycemia were previously known to induce structural changes in the lungs, giving rise to pulmonary remodelling and the subsequent restrictive respiratory pattern.^47^ Moreover, hyperglycemia is also known to generate reactive oxygen species and induces oxidative stress^44^, leading to endothelial dysfunction which may cause further hyperglycemic pulmonary microangiopathy.^48^ This is in line with the findings of a study by Lampasona et al. which demonstrated that inflammation and coagulopathy, rather than impaired antibody response as such present in individuals with diabetes, were more responsible in aggravating the outcomes.^49^ Therefore, this explains the poorer prognosis found in hyperglycemic patients without prior diabetic history, and again supporting the direct link between glucose level and disease progression.

Altogether, these findings illustrate the potential utility of admission BG as a predictor for poor prognosis in COVID-19 patients. Considering that BG measurement is relatively practical and instant, its quantification upon admission would be beneficial in predicting the likelihood of progression to severe COVID-19 cases. Therefore, we encourage clinicians to routinely obtain FBG values of each COVID-19 suspected case at admission, thus providing a simple method of risk stratification for management of patients in clinical settings, which would be particularly helpful in streamlining the limited number of medical resources during the current pandemic. Although our findings also favored over the use of admission RBG, future researches are required as we were unable to comprehensively explore the independent prognostic value of admission RBG due to paucity of studies. Furthermore, the current evidence indicated that the cut-off values to predict poor prognosis in COVID-19 patients are still equivocal^31,34,50,51^, suggesting that future large multicenter studies are required to obtain the most optimum cut-off value.

Despite the fact that our findings favored over the prognostic value of high admission BG, the observed unexplained heterogeneity and the fact that all studies were retrospective and conducted in China may limit the generalizability of our findings. Furthermore, most of the studies included in the DRMA on severity outcome yielded moderate risk of bias, thus indicating potential overestimation of the observed effects due to imprecision. These indicated that the observed effects should be interpreted cautiously, and future studies with higher quality of evidence are required to confirm the estimated risks. Nonetheless, our results were consistent with the independent prognostic value of admission FBG, thus we judged the certainty of evidence for severity to be high, and for mortality and poor outcome to be moderate. To the best of our knowledge, this is the first meta-analysis conducted to show the potential use of admission BG as a predictor of poor prognosis in COVID-19 patients. Although our eligibility criteria may introduce language bias, our study included a relatively large number of cohorts and only four non-English articles were excluded^52–55^, suggesting that any potential bias was negligible. We hope that our findings may enhance the current knowledge on the management of COVID-19, thus contributing to the alleviation of the devastating disease burden.

## CONCLUSION

In conclusion, this meta-analysis adds to the growing body of evidence corroborating the potential utility of admission BG as a predictor of poor prognosis in COVID-19 patients. There is a high-quality evidence on the prognostic value of admission FBG towards severity, and moderate-quality evidence on its prognostic value towards mortality and poor outcome, while the other outcomes yielded very low-to-low quality evidence. In addition, we demonstrated non-linear dose-response relationship between admission FBG and COVID-19 severity. Further studies to ascertain the estimated risk in the DRMA and to confirm the observed prognostic value of admission RBG are required.

## Data Availability

The data that supports the findings of this manuscript is available within the supplementary files.

## ACKNOWLEDGEMENT

The authors would like to express their gratitude to Dr. Long Qi (Department of Critical Care Medicine, Hubei Provincial Hospital of Traditional Chinese Medicine, China), Dr. Ting Chen (Department of Endocrinology, Union Hospital, Tongji Medical College, Huazhong University of Science and Technology, China), as well as Prof. Gian Paolo Fadini and Dr. Mario Luca Morieri (Department of Medicine, University of Padova, Italy) for the provision of additional data for analysis. The authors would also like to thank Dr. Miguel Marcos (Department of Internal Medicine, University Hospital of Salamanca-IBSAL, University of Salamanca, Spain), Dr. Celestino Sardu (Department of Advanced Medical and Surgical Sciences, University of Campania “Luigi Vanvitelli,”, Italy), Prof. Giuseppe Penno (Section of Diabetes and Metabolic Diseases, University of Pisa, Italy) and Dr. Juan Berenguer (Hospital General Universitario Gregorio Marañón, Spain) for the confirmation of study settings. Lastly, the authors would like to acknowledge Dr. Eka Dian Safitri (Clinical Epidemiology and Evidence-Based Medicine Unit, Dr. Cipto Mangunkusumo General Hospital - Faculty of Medicine Universitas Indonesia, Jakarta, Indonesia) and Dr. Besral (Department of Biostatistics, School of Public Health University of Indonesia, Depok, West Java, Indonesia) for the methodological and statistical advices.

## AUTHOR CONTRIBUTIONS

GL and DLT conceptualized the idea for the project and designed the methodology. GL, JA, VKW, and AT performed the literature search, study screening, and data abstraction. GL administered the study protocol, undertook the formal analysis, and visualized the results. GL, JA, and VKW developed the risk of bias tool and drafted the manuscript, and risk of bias assessment was conducted by JA and VKW. GL, VKW, AT, and DLT reviewed and edited the manuscript for final submission. DLT validated and supervised the project. All authors have approved of the final manuscript for publication.

## CONFLICT OF INTERESTS

The authors declare no conflict of interest.

## FUNDING

This project received no specific grant from any funding agency in the public, commercial, or not-for-profit sectors.

## Notes

### Competing Interest Statement

The authors have declared no competing interest.

### Clinical Protocols

https://www.crd.york.ac.uk/PROSPERO/display_record.php?RecordID=202224

